# Development of an IVF prediction model for donor oocytes: a retrospective analysis of 9,384 embryo transfers

**DOI:** 10.1101/2024.04.04.24305303

**Authors:** Oisin Fitzgerald, Jade Newman, Luk Rombauts, Alex Polyakov, Georgina M Chambers

## Abstract

**Study question:** Can we develop a prediction model for the chance of a live birth following transfer of an embryo created using donated oocytes?

**Summary answer:** Three primary models that included patient, past treatment and cycle characteristics were developed to predict the chance of a live birth following transfer of an embryo created using donated oocytes; these models were well calibrated to the population studied and achieved reasonable predictive power.

**What is known already:** Nearly 9% of assisted reproductive technology (ART) embryo transfer cycles performed globally use embryos created using donated oocytes. This percentage rises to one quarter and one half in same-sex couples and women aged over 45 years respectively.

**Study design, size, duration:** This study uses population-based Australian clinical registry data comprising 9,384 embryo transfer cycles that occurred between 2015 and 2021.

**Participants/materials, setting, methods:** Three prediction models were compared that incorporated patient characteristics, but differed in whether they considered use of prior autologous treatment factors and current treatment parameters. We evaluated the models using grouped cross validation and report several measures of model discrimination and calibration. Variable importance was measured through calculating the change in predictive performance that resulted from variable permutation.

**Main results and the role of chance:** The best performing model has an AUC-ROC of 0.60 and Brier score of 0.20. While this indicates approximately 15% less discriminatory ability compared to models assessed on an autologous cohort from the same population the performance of the models was clearly statistically significantly better than random and well calibrated to the population studied. The most important variables for predicting the chance of a live birth were the oocyte donor age, number of prior oocyte recipient embryo transfer cycles and whether the transferred embryo was cleavage or blastocyst stage. Of lessor importance were the oocyte recipient parity, whether donor or partner sperm was used, the number of prior autologous embryo transfer cycles and the number of embryos transferred.

**Limitations, reasons for caution:** The variation in donor oocyte cohorts across countries due to differences in whether anonymous and compensated donation are allowed may necessitate the models be re-calibrated prior to application in non-Australian cohorts.

**Wider implications of the findings:** These results confirm the well-established importance of oocyte age and ART treatment history as the key prognostic factors in predicting treatment outcomes. One of the developed models has been incorporated into a consumer-facing website (YourIVFSuccess.com.au/Estimator) to allow patients to obtain personalised estimates of their chance of success using donor oocytes.

**Study funding/competing interest(s):** This research was funded by the Australian government as part of the Medical Research Future Fund (MRFF) Emerging Priorities and Consumer Driven Research initiative: EPCD000007.

**Trial registration number:** N/A

## Introduction

Each year there are more than 3 million assisted reproductive technology (ART) embryo transfer cycles worldwide (Adamson, 2022). Of these, nearly 9% use embryos created using oocytes donated to the intending mother by an anonymous or known donor. This percentage increases markedly in women over 45; in this age group donated oocytes account for one third of Australian ART cycles (Newman et al., 2022), and more than 50% of US ART cycles (Klein & Sauer, 2012). Oocyte ‘donation’ also occurs at a higher rate in same-sex female couples with one third involving use of donor oocytes (Newman et al., 2022), in many cases as provision of oocytes from one partner to the other (Bodri et al., 2018). While several prediction models have been developed using clinical registries for the purpose of counselling women undertaking ART on their chance of a successful live birth (Leushuis et al., 2009; Nelson & Lawlor, 2011; Ratna et al., 2020), these are often exclusively been based on data where women used their own oocytes/embryos (autologous cycles) or have unclear discriminatory ability on oocyte recipient cycles. Further, in cases where oocyte donation cycles are included (Luke et al., 2014), jurisdictional differences in oocyte donation programs may mean the predicted success rates fail to generalise across countries. This, coupled with the tendency for traditionally marginalised groups such as same-sex couples to be over-represented in the use of oocyte donation cycles (Newman et al., 2022) creates a need for the development and/or evaluation of donor oocyte based prediction models.

(Ratna et al., 2020). ART prediction models when made available to consumers as personalised calculators for estimating treatment success, help patients feel more comfortable with the decision-making process and improve communication between patients and clinicians (Stacey et al., 2017). The Australian government funded *YourIVFSuccess* website was launched in 2021, and includes a *Patient Estimator* to predicted the patients chance of a live birth based on their individual characteristics and past fertility treatment history (YourIVFSuccess, 2023). A mixed-methods evaluation study of the website found that the Patient Estimator aided in realigning patient expectations in approximately 74% of cases, with the majority of study participants finding it to be trustworthy and helpful (Brew et al., 2023). The main criticism of the Estimator was limited or unclear applicability to treatment cycles involving donor gametes, single women and same-sex couples.

Variation in legal and regulatory systems characterised by whether anonymous donation is allowed (Harper et al., 2016), the existence of egg-sharing programmes (Ahuja et al., 1996), whether the donor can be reimbursed (Kenney & McGowan, 2014), and the existence of government subsidies for the process (Rabinerson et al., 2002) may impact the prognostic characteristics of the local donor population. This creates a need to ensuring donor-based prediction models are well-calibrated to a population of interest rather than naïve application of a model developed on a different population. In Australia federal law prohibits payment for oocyte donation beyond “reasonable expenses incurred” (Commonwealth of Australia, 2002; Goedeke et al., 2020), with anonymous donation prohibited legally in three states, and across the country by the peak medical regulatory body (NMHRC, 2023). These factors may influence willingness or ability to donate. For instance, compared to a cohort of 1,423 donors from 11 European countries (Pennings et al., 2014), the population of Australian donors tend to be older (32 vs 27 years) (Harris et al., 2016) an important prognostic factor in predicting the chance of a successful live birth.

Previous research has investigated the association between patient level factors such as the age, BMI, infertility diagnoses and ethnicity of the donors and recipient, along with treatment characteristics on oocyte recipient success rates. Oocyte donor age has consistently been shown to be an important factor (Cohen et al., 1999; Hogan et al., 2019; Wang et al., 2012), with a non-linear association with the chance of a live birth analogous to that seen for autologous ART cycles, with a drop in success rates around 35 years that is exponential as age increases. In contrast, the impact of recipient age is largely nullified, with a decrease in success rates observed at 45 years (Soares et al., 2005) or not at all (Pataia et al., 2021; Wang et al., 2012). Other pretreatment factors demonstrating evidence of an impact on recipient cycle outcomes include recipient ethnicity (Zhou et al., 2020), endometrial thickness (Noyes et al., 2001), and paternal age (Begon et al., 2023) with mixed evidence for the impact of donor and recipient BMI (Cardozo et al., 2016; Jungheim et al., 2013; Pataia et al., 2021; Xu et al., 2021). Treatment factors or intermediate outcomes such as the stage of embryo development also impact the chance of a successful outcome (Kontopoulos et al., 2019). However, the degree to which these patient and treatment factors can accurately predict oocyte recipient cycle live birth rates is unclear, with a lack of research following best practices in clinical prediction tool development (Lee et al., 2016; Ratna et al., 2020) in assessing the effectiveness of models in predicting the outcome of oocyte recipient cycles. Understanding a models strengths and weaknesses is important information in cases where these models may be used as decision aids for informing prospective ART patients on their chance of success, given evidence that providing such information can result in altered clinical practice and patient behaviour (Brew et al., 2023).

In summary, there is extensive variation in the practice and legality of donor oocyte-based ART across the world. This increases the need for local development and/or evaluation of clinical prediction models for this purpose. Further, few past papers have reported predictive validation metrics for oocyte donor models making it unclear how they are likely to perform. The goal of this article is to use Australian clinical registry data to develop and compare models that predict the likelihood of a live birth after an embryo transfer using donated oocytes. This information will aid in informing those individuals considering the use of donated oocyte ART on their chance of a live birth following treatment.

## Methods

### Cohort

Data for this project were sourced from the Australia and New Zealand Assisted Reproduction Database (ANZARD), a Clinical Quality Registry comprising information on all assisted reproductive technology (ART) treatment cycles undertaken in Australian and New Zealand fertility clinics. It is a requirement of an ART Unit’s (IVF clinic’s) licence to submit data to ANZARD, thus complete ascertainment is assumed. From ANZARD we extracted a cohort consisting of 4,853 women (recipients) who underwent their first oocyte recipient embryo transfer cycle in Australia between 2015 and 2021. Following exclusion of cycles that related to an attempt to achieve a second or subsequent children using donor oocytes and exclusion of 89 cycles missing data our final cohort consisted of 9,384 embryo transfer cycles involving 4,811 women (recipients) receiving treatment at 84 IVF clinics. See Supplementary Figure A1 for a more detailed characterisation of the impact of the study inclusion and exclusion criteria on the final cohort. We extracted information on the number of past autologous treatment cycles (if any), the current recipient embryo transfer number, oocyte recipient age, the oocyte donor age, the sperm provider (paternal) age, the parity of the recipient, whether pre-implantation genetic testing (PGT) was performed and whether the transfer was a single or double embryo transfer of fresh or thawed embryo at cleavage or blastocyst stage. The study outcome, live birth, was defined as the birth of at least one live infant of at least 20 weeks gestation or a minimum of 400 grams birthweight.

### Analysis

#### Descriptive analysis

The demographic and treatment characteristics of the analysis cohort are described using counts (%) for categorical variables, and medians (25^th^ percentile (Q1) and 75^th^ percentile (Q3))) for continuous variables stratified by whether a live birth occurred. A *t*-test (continuous variable) or *chi-*squared test (categorical variables) was used to assess whether the univariate distribution differed between those who achieved a live birth and those who did not for each variable. Additionally, for each of the three age variables (recipient age, oocyte donor age, sperm provider age) we graphically characterise its distributional spread and marginal relationship with live birth rate.

#### Statistical models

The aim of the statistical modelling was to assess which factors best predicted a live birth following transfer of a donor oocyte embryo. Live birth was coded as a binary variable (1=live birth; 0=no live birth) with the models predicting the probability of this event. The following variables were used in construction of prediction models: number of previous autologous embryo transfers (coded as 0, 1-3 and 4+), oocyte recipient age at embryo transfer (years), oocyte donor age at donation (years), sperm provider age at embryo transfer for fresh freezing for frozen-thaw transfers (years), oocyte recipient parity (1=parous; 0=nulliparous), whether this was the recipient’s first, second or third or more donor oocyte embryo transfer, whether the transfer was of a fresh or thawed embryo, the number of embryos transferred, the stage of embryo development of the transferred embryos, and whether ICSI was used to fertilise the transferred embryos.

We assessed the performance of 3 models at the prediction task. These pooled generalised additive models (GAMs) summarised below, varied in whether they considered prior autologous treatment characteristics (if any) and the current embryo transfer characteristics as input features. GAMs are extensions of generalised linear models (GLM) (in this case logistic regression) that allow for adaptive modelling of non-linear responses using penalised spline transformation of continuous variables, with the use of a spline transformation indicated using f(.) below (Hastie & Tibshirani, 1995).

**Model 1:** Live birth ∼ donor embryo transfer number + f(oocyte recipient age) + f(oocyte donor age) + use of partner sperm + use of male partner’s sperm * male partner’s age + nulliparous

**Model 2:** Model 1 + number of prior autologous embryo transfers

**Model 3:** Model 2 + transfer of two or more embryos + use of frozen embryo(s) + use of IVF/ICSI embryo(s) + transfer of cleavage/blastocyst stage embryo(s)

Unlike oocyte recipient and donor age, male partner age was not modelled using a spline transformation as in prior analysis and research, male partner age was not found to be significant. Further, sperm provider age only appears in the interaction term “use of male partner’s sperm * male partners age” as the source database does not collect sperm provider age for donor sperm cycles.

#### Predictive performance

The predictive performance of the models was assessed using several metrics: the area under the receiving operating curve (AUC-ROC or c-statistic) (optimal=1.0), the area under the precision recall curve (AUR-PR) (optimal=1.0), the Brier score (or mean squared error) (optimal=0.0) and the Brier skill score (optimal=1.0). The AUC-ROC assesses the discrimination power of the model, i.e., whether the model tends to give higher probabilities to positive cases (those where a live birth occurs) than negative cases (those where a live birth does not occur), the AUC-PR is a measure of how confidently the models detects positive cases (does it give them high probabilities), and the Brier score is a general measure of both the prediction calibration and certainty. The Brier skill score compares the Brier score of a proposed model with a reference model, in this case we use the population level chance of a live birth per embryo transfer of 28% (Table 1). Calibration is whether the predicted rate of occurrence (e.g., a 10% chance of a live birth) matches the observed rate (e.g., a live birth rate of 10%). Along with the Brier score, calibration was directly assessed by splitting each model’s predicted probabilities into 5 bins of roughly equal size (i.e., the quantiles) and assessing graphically whether each bin’s average predicted probability of a live birth matched the observed rate.

**Table 1.**
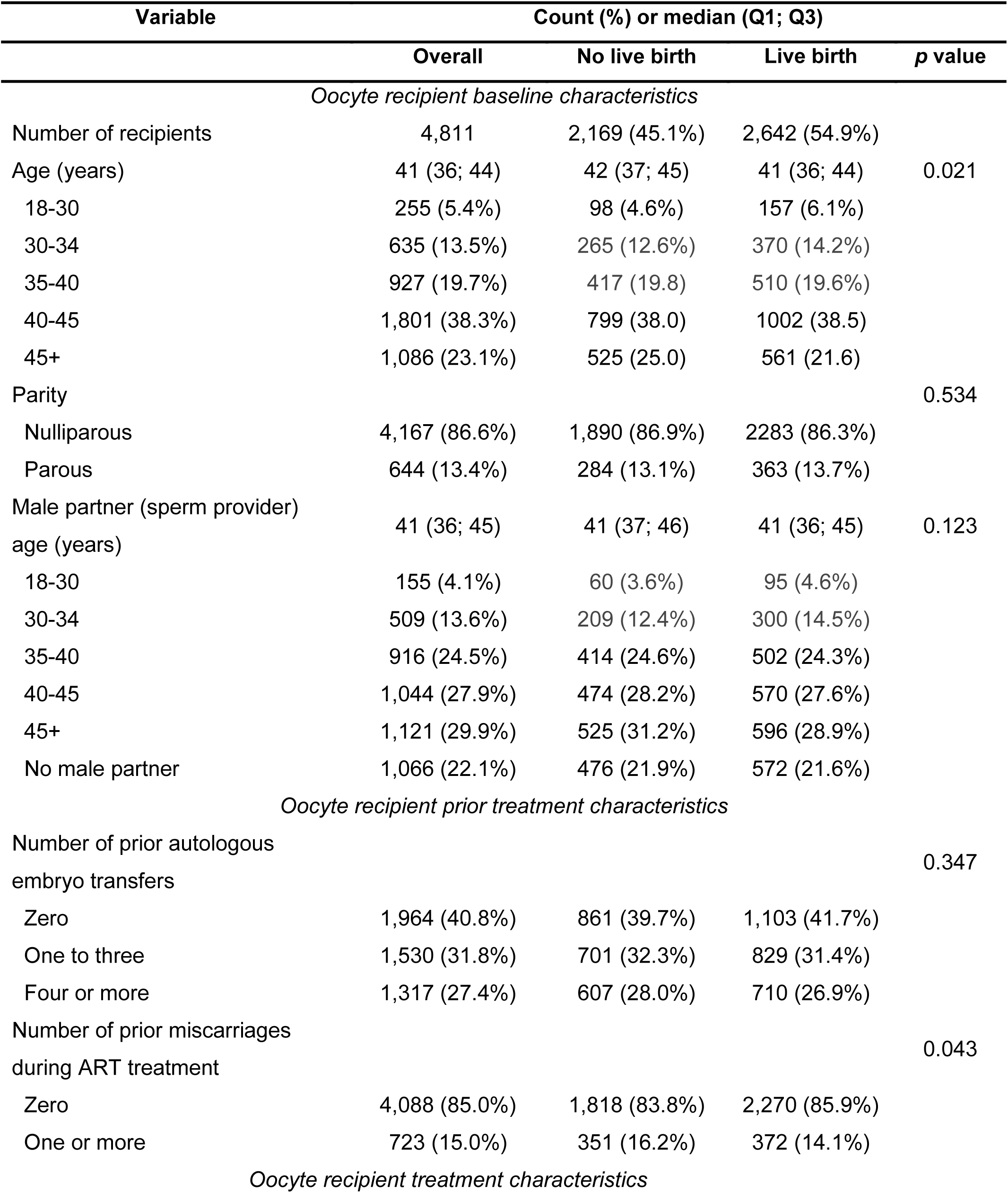
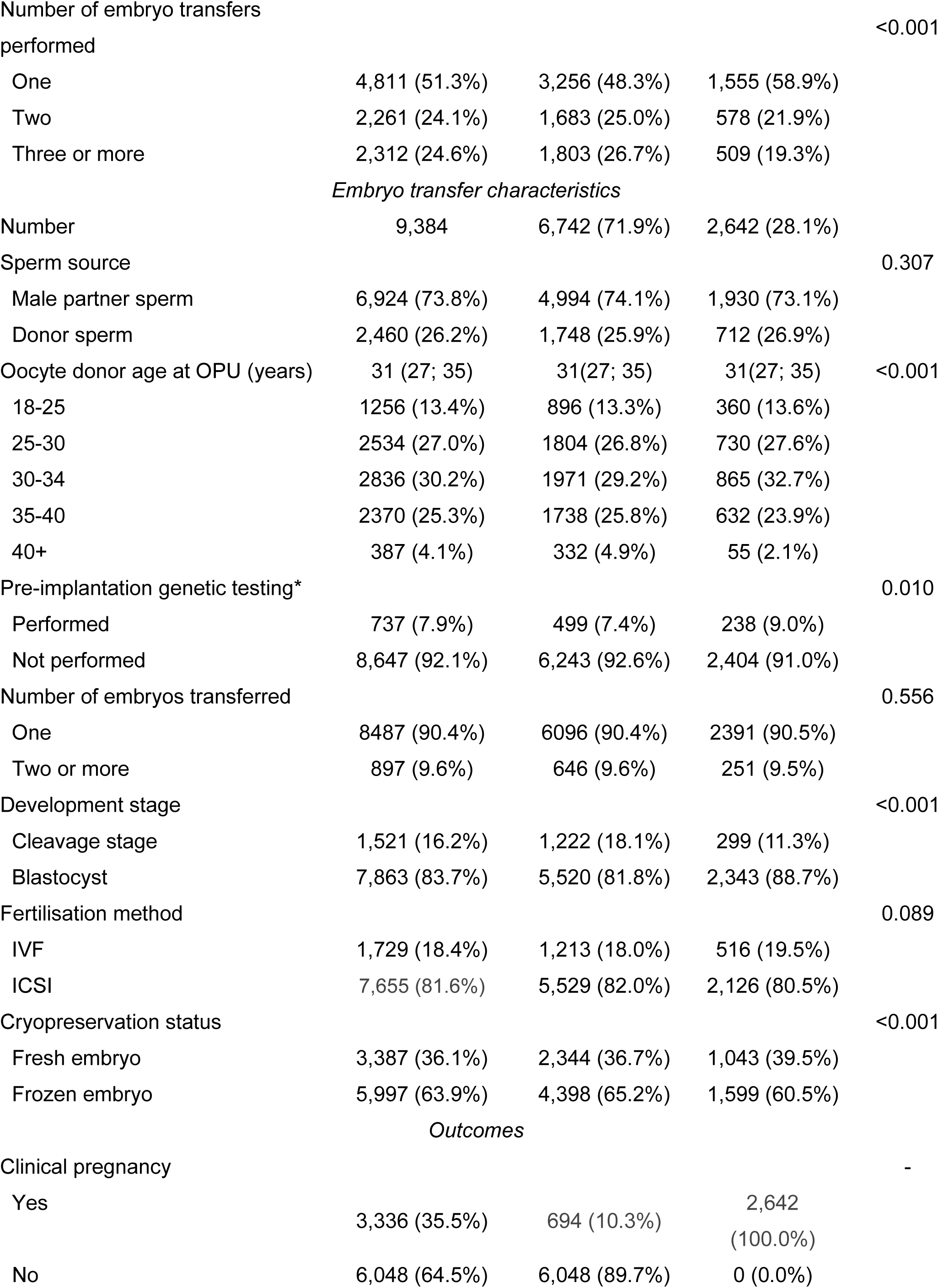
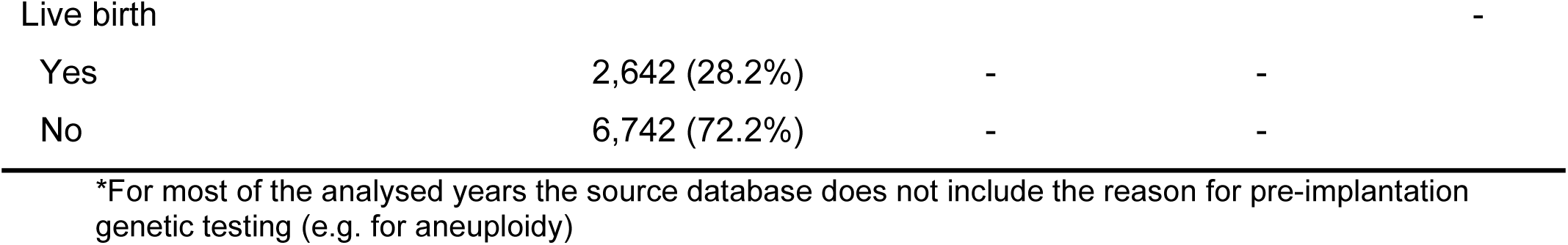
Characterisation of study cohort used to develop an IVF prediction model for donor oocytes. Data sourced from ANZARD (2015-2021).

These metrics were calculated using grouped 10-fold cross-validation. This involves repeatedly splitting the data into a portion containing 90% of women (recipients) to use in training the models and leaving a 10% test sample for calculation of the model performance metrics. This entire process was then repeated 10 times to account for variability introduced by the data splitting process, with post-hoc pairwise comparisons of the models (α=0.05) across each metric performed using a *t* statistic with the variance corrected to account for the non-independence across fold and repetitions (Bouckaert & Frank, 2004). More detailed assessment (calibration and calculation of performance metrics) of the top performing models was also performed stratifying by age variables and treatment factors.

#### Variable predictive importance

The predictive importance of the variables in the models was assessed using permutation feature importance (Breiman, 2001). This algorithm involves variable by variable random shuffling of the values, and calculation of the above performance metrics after making a prediction using the shuffled dataset. For variables of high importance, a large degradation in performance should be observed, while little impact should be seen for unimportant variables. While the model coefficients and their standard errors also provide this information, we present this as complimentary information given coefficients and their standard errors may be overfit to the training data.

We report the importance measure as the percentage change in performance observed compared to the baseline model in the relevant cross-validation fold and compare, for each variable, whether this change in performance is different from zero using the Wilcoxon signed rank test (α=0.05). While this approach may lead to a higher Type 1 (false positive) error rate than the corrected *t-*test used above (Bouckaert & Frank, 2004), we believe mistakenly including an unimportant variable is preferrable to mistakenly leaving out an important variable.

#### Model coefficients and interpretation

The results of the best performing models, fitted to the entire dataset, are presented as a mix of the model coefficients (on the odds scale) for binary and categorical variables along with graphical display of the spline effects.

### Ethics and reporting guidelines

Ethics for this project was obtained from the UNSW Sydney Human Research Ethics Committee (reference number: iRECS0859). We followed the TRIPOD reporting guidelines for development of a prediction model (Collins et al., 2015).

## Data Availability

The data underlying this article cannot be shared publicly due to patient privacy concerns.

## Funding

This research was funded by the Australian government as part of the Medical Research Future Fund (MRFF) Emerging Priorities and Consumer Driven Research initiative: EPCD000007.

## Data availability statement

The data underlying this article cannot be shared publicly due to patient privacy concerns.

## Results

### Descriptive statistics

As shown in Table 1 the study cohort of oocyte recipients were older than the population of women who undertake autologous ART cycles reported in previous research (Chambers et al., 2017). While around 11% of first-time autologous cycles are in women over 40 years, in recipient cycles this rose to over 60% (Table 1, Figure 1A and 1B). Of course, oocyte recipients are not necessarily ART naïve and in this cohort 59.2% had undertaken prior autologous cycles, with the autologous cycles largely unsuccessful. The vast majority (86.6%) of oocyte recipient were nulliparous (Table 1). Summarising the average oocyte recipient, they were 41 years old, nulliparous, around 60% had previously done ART and had a male partner aged 41 years (Table 1 and Figure 1C). As expected, the oocyte donors were a younger population, and were on average 31.0 (median 31) years, with the majority aged between 25 and 35 years (Table 1 and Figure 1A). The treatment characteristics of the study cohort was similar to the general ART population in Australia (Newman et al., 2022), where 90.4% of cycles were single embryo transfers (SET), 83.7% of cycles involved a blastocyst transfer, and 63.9% of cycles were performed as frozen-thawed embryo transfer cycles (Table 1), although this rate was far lower on the first donor embryo transfer (40.3%)

**Figure 1.**
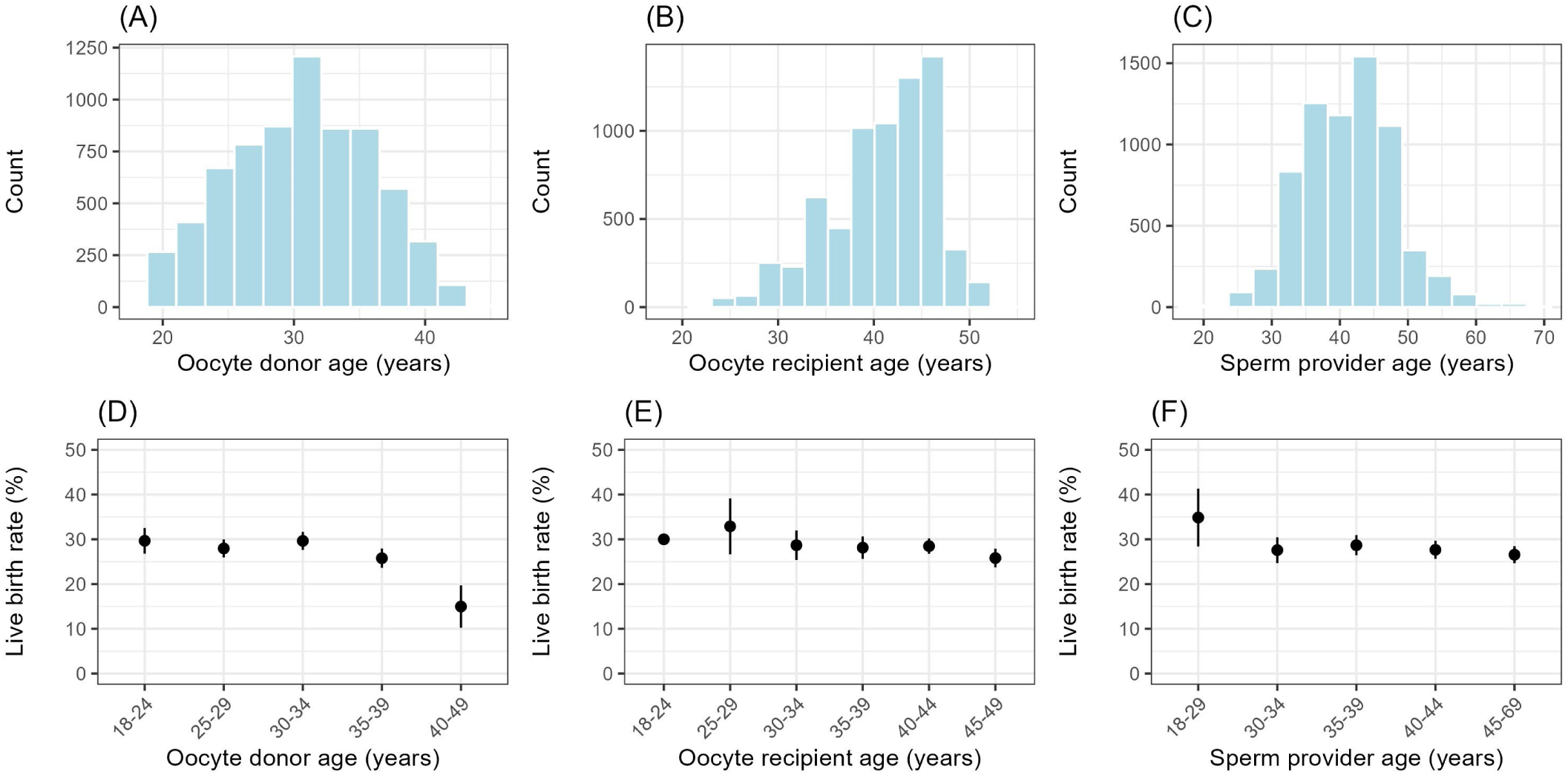
Characterisation of age variables used in development of an IVF prediction model for donor oocytes. Distribution of and live birth rate by oocyte donor age (A and D), oocyte recipient age (B and E) and sperm provider age (C and F). Data sourced from ANZARD (2015-2021).

Figure 1D-F shows live birth rates by the three age variables (oocyte donor age, oocyte recipient age and sperm provider age). Increasing oocyte donor age had a clear negative impact on the marginal live birth rates. From Figure 1E we see a decrease in the live birth rate from around 35 years, matching patterns found in previous research. In contrast, recipient age had a less pronounced decrease that only occurred around 45 years. Sperm provider age appearing to have little relation with live birth rate (Figure 1F).

### Predictive performance

As shown in Table 2 the top performing model was Model 3, this uses the most input variables and has the form:

**Table 2.** Cross-validation results from development of an IVF prediction model for donor oocytes using ANZARD (2015-2021). Outcome of the 10-times 10-fold grouped (by patient) cross validation model evaluation reported as metric mean (standard error). The best and any statistically indistinguishable (α=0.05) metrics are bolded.

| Model | Brier score | Brier skill score | AUC ROC | AUC PR |
| --- | --- | --- | --- | --- |
| 1 | 0.1991 (0.0070) | 0.0162 (0.0066) | 0.5811 (0.0203) | 0.3301 (0.0222) |
| 2 | 0.1991 (0.0070) | 0.0162 (0.0067) | 0.5816 (0.0205) | 0.3308 (0.0229) |
| 3 | **0.1978 (0.0069)** | **0.0230 (0.0066)** | **0.5970 (0.0198)** | **0.3422 (0.0237)** |

Live birth ∼ donor embryo transfer number + f(oocyte recipient age) + f(oocyte donor age) + use of partner sperm + use of male partner’s sperm * male partner’s age + nulliparous + number of prior autologous embryo transfers transfer of two or more embryos + use of frozen embryo(s) + use of IVF/ICSI embryo(s) + transfer of cleavage/blastocyst stage embryo(s)

Comparison of the model metrics using paired *t*-tests demonstrated that the performance of Model 3 is statistically significantly (*α*=0.05) better than Models 1 and 2 across all metrics (*p*<0.01). There was no statistically significant difference between Models 1 and 2, except for the AUC-PR (*p*<.01) where Model 2 was marginally superior. The discriminatory ability of Model 3 is displayed graphically in Figure 2A where we see significant overlap in the predicted probabilities. Figures 2B-D illustrate the model average performance (in this case the Brier score) stratified by oocyte donor age, oocyte recipient age and year of treatment, with only oocyte donor age illustrating any association with model performance. From Figure 2E we see the predictions from Model 3 are well calibrated with little deviation between the predicted vs. observed live birth rate across the bins. These findings were similar across Models 1 and 2.

**Figure 2.**
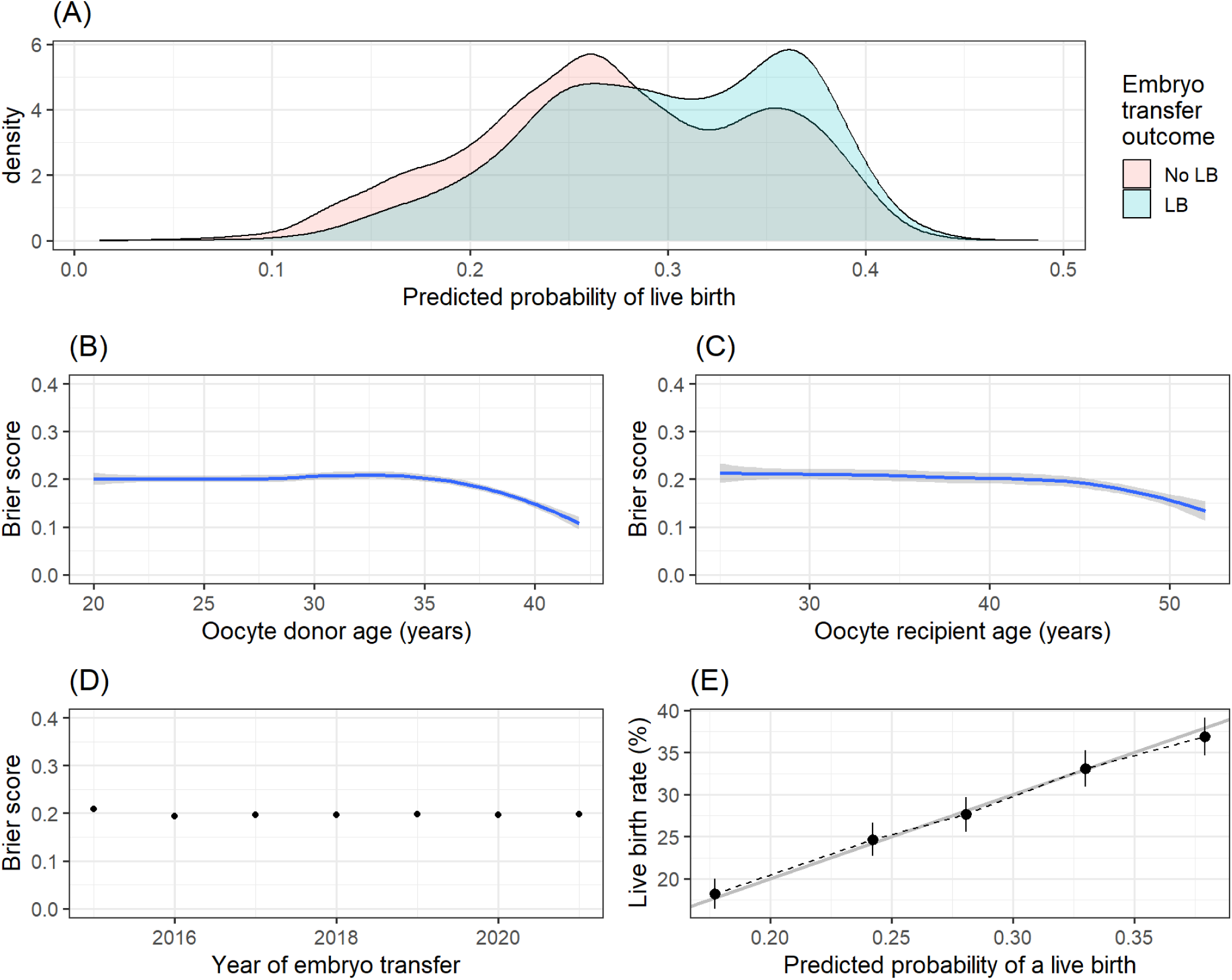
Results from development of an IVF prediction model for donor oocytes using ANZARD (2015-2021). Stratified analysis of the performance of Model 3. (A) Graphical illustration of model discriminatory power. Smoothed estimated of Brier score by (B) oocyte donor age, (C) oocyte recipient age and (D) year of embryo transfer. (E) Predicted probability calibration curve.

### Variable predictive importance

As the best performing model, the variable importance measures for Model 3 can be found in Figure 3. Generally, few of the variables considered had a clear impact on model predictive ability as illustrated by clear model performance degradation (e.g. decreased AUC-ROC) under permutation of the variable. These included oocyte donor age (% decrease in AUC-ROC=3.7, *p*<0.01), oocyte recipient age (% decrease in AUC-ROC=1.0%, *p*=0.02), number of prior donor oocyte transfers (% decrease in AUC-ROC=6.3, *p*<0.01) and whether the transferred embryo was cleavage or blastocyst stage (% decrease in AUC-ROC=4.9, *p*<0.01) of clear importance for model performance (Figure 3). Variables with of lessor importance included parity (% decrease in AUC-ROC=0.5, *p*<0.01), use of partner or donor sperm (% decrease in AUC-ROC=0.2, *p*<0.01), number of embryos transferred (% decrease in AUC-ROC=0.1, *p*=0.04), number of prior autologous embryo transfers (% decrease in AUC-ROC=0.2, *p*<0.01). Paternal (sperm) age, use of fresh or frozen-thawed embryos and use of ICSI did not appear to impact predictive ability.

**Figure 3.**
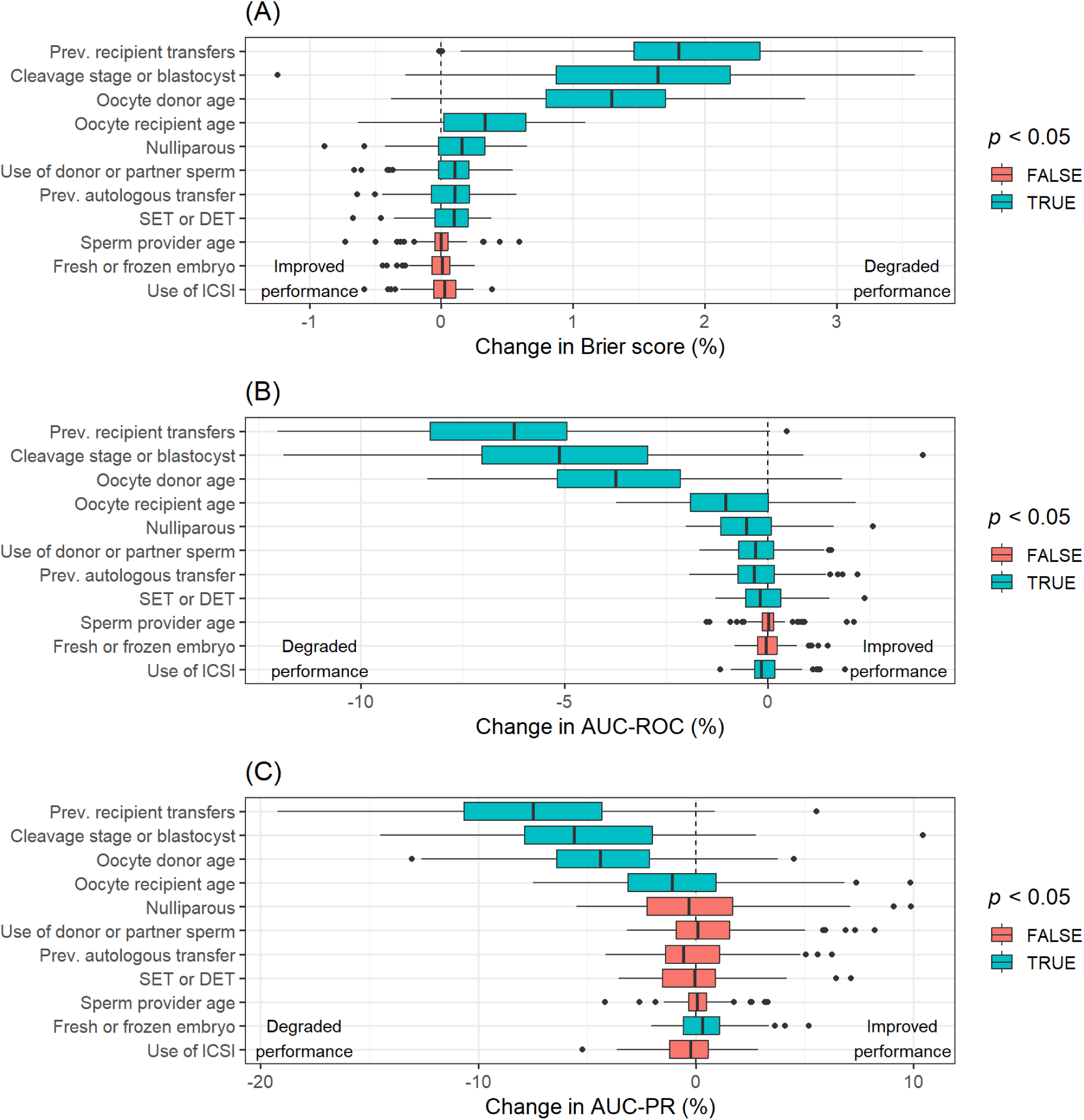
Results from development of an IVF prediction model for donor oocytes using ANZARD (2015-2021). Permutation feature importance analysis results for Model 3, with colouring to indicate whether a hypothesis test that the change in the metric was centred around zero (Wilcoxon signed rank test) was significant at α=0.05. The boxplots display the distribution of the percentage change in the (A) the Brier score (an increase indicates a degradation in performance), (B) the AUC-ROC (a decrease indicates a degradation in performance), and (C) the AUC-PR (a decrease indicates a degradation in performance) that results from permuting each feature across the 10-times 10-fold cross-validation. The further the distribution of the percentage change in the metric is from zero, the greater the degradation in model performance as a result of permuting that variable.

### Model coefficients and interpretation

Graphical and tabular display of the spline and parametric coefficients for Model 3 are found in Supplementary Figure A2 and Supplementary Table A1. We modelled oocyte donor and recipient ages as non-linear effects. However, the non-linearity of donor age was far more pronounced, demonstrating the prescribed mid-thirties age related fall in the chance of a live birth (Supplementary Figure A2). The parametric coefficients for Model 3 have interpretations in line with expectations. For Model 3 - an increased odds of a live birth for parous women of 22% (*p*=0.07), a reduction in the odds of a live birth of 12% (*p*=0.04) for women who have undergone more than 4 prior autologous embryo transfers (compared to none), a reduction in the odds of a live birth with each additional attempt by 20-30% (*p*<0.01), a reduced odds of a live birth by 44% (*p*<0.01) when a cleavage stage embryo is transferred and an increased odds of a live birth by 17% (*p*=0.04) when more than one embryo is transferred. Male partner age, source of sperm (donor or husband), use of IVF/ICSI and transfer of a fresh or thaw embryo were all clearly non-significant. Aside from sperm source, the coefficient standard errors and *p*-values are largely in line with the variable importance results (Figure 3), in that finding of variable importance implied a more precise coefficient estimate (e.g., *p*-value <0.05).

## Discussion

The goal of this study was to develop and compare clinical prediction models for the likelihood of a live birth after an ART embryo transfer using donated oocytes for the purpose of informing the development of the YourIVFSuccess.com.au website. The models were developed on 9,384 embryo transfers performed in Australia between 2015 and 2021. While the discriminatory power of the models was quite weak, with AUC-ROC values of 0.58 and 0.60 depending on whether treatment factors were considered (Table 2), the predictions appeared well calibrated (Figure 2E) providing useful information on the chance of a live birth that can be tailored to the individual where additional prognostic information is available. We found that oocyte donor age followed by recipient age and parity were the most important pre-treatment factor in predicting a live birth, with sperm provider age having limited or no contribution to forecasting a live birth (Figure 3). When treatment factors were considered, the number of prior donor oocyte embryo transfers and the stage of embryo development at transfer improved model discriminatory power by 6.3% and 4.9% respectively, while number of embryos transferred, use of donor or partner sperm, and number of prior autologous cycles were associated with lessor improvements of 0.1-0.2%. On the other hand, embryo cryopreservation status and fertilisation method had no impact on predictive power (Figure 3).

Variation in laws, regulations and cultural beliefs around the use of donor gametes mean that oocyte donation and recipient cohorts may vary across jurisdictions, possibly more so than autologous ART cohorts, necessitating care and appropriate recalibration before applying models across jurisdictions. Nevertheless, the cohort making up the current research share some characteristics with previously reported oocyte recipient cohorts. In line with a large (N=9,865) US based study (Kawwass et al., 2013) the oocyte recipients are typically over 40 years, with 23.1% over 45. While the data source does not collect the reason for use of donor oocytes, nearly two-thirds (59.1%) of the current cohort had done at least one previous autologous embryo transfer, only slightly higher than the US cohort (54.5%) (Kawwass et al., 2013), suggesting recurrent IVF failure (possibly in combination with age) as the biggest drivers for use of donor oocytes in these jurisdictions. In comparison, a Belgian based study of 144 couples found that 38.9% had done previous ART (68.8% any previous fertility treatment) and 41.7% were using donor oocytes due to age or recurrent failure (Baetens et al., 2000).

In contrast to US studies which have reported average donor ages of 26 years (Roeca et al., 2020), but as previously observed (Wang et al., 2012), the Australian oocyte donors were older, with a median (average) age of 31 (30.6) years, largely attributable to prohibitions on payment for donation and anonymous donation (Goedeke et al., 2020; NMHRC, 2023).

However, as few (4.1%) recipient embryo transfers used a donor aged 40 or over, most oocyte recipient embryo transfers had a predicted live birth rate per embryo transfer of approximately 30% (Table 1 and Figure 1D). A question for future research is whether other characteristics of Australian oocyte donors (or recipients) which potentially impact ART success rates such as BMI (Cardozo et al., 2016) are different, given US oocyte recipients appear to have a higher live birth rate per embryo transfer at nearly 50% (Roeca et al., 2020) or whether this difference may also be explained by variation in treatment. For instance, US recipient cycles are more than twice as likely to involve multiple embryo transfer and up to four times as likely to incorporate PGT compared to Australian (Roeca et al., 2020; SART, 2024).

The current models had lower discriminatory power than that reported for most predictive models based on autologous cohorts. For instance, in the current research the AUC-ROC is at best 0.6 (using treatment information), in contrast to autologous (post-treatment) complete cycle models, which typically score closer to 0.7 (Ratna et al., 2020). A key reason for this difference is less variation in the age of oocyte donors, with only 4.1% over 40 years, in contrast to autologous cycles where nearly 30% of women are over 40 (Newman et al., 2022). As shown in Figure 2B for the Brier score, predictive performance improves when the oocyte donor is over 40 years due to increased certainty about the likely poor outcome. As a result, lower numbers of oocyte donors in this age group are likely a key explanation for the difference in AUC-ROC metrics between donor and autologous models rather than donor cycle outcomes being inherently more difficult to predict. Indeed, arguably it is at the upper limits of oocyte donor and recipient age that such models are likely to have greatest value in setting patient expectations (Brew et al., 2023). Given even autologous models that include detailed patient history and physiological measures fail to increase the AUC-ROC by more than 10% (to 0.77) (Ratna et al., 2020), these results highlight the degree to which there is large uncertainty about the likely the chance of ART success, and the extent to which ART treatment history is a key prognostic factor. As shown in Figure 3 the number of prior recipient embryo transfers has a relatively large impact on the model performance, with such repeated failures acting a proxy for underlying conditions not recorded in the data (Rozen et al., 2021).

The feature importance findings of the current study are largely in line with previous research. Oocyte donor age was the most important non-treatment factor in predicting the probability of a live birth (Figure 3), with a partial effect (Supplementary Figure A2) similar to the effect observed for oocyte age in autologous cycles (e.g., Figure 3 of Newman et al. (2022)). While we did find evidence of a post 45-years decline in the oocyte recipient chance of success (Figure 2C and Supplementary Figure A2), similar to previous research (Soares et al., 2005), this effect was relatively weak and the predictive importance of oocyte recipient age was approximately one-quarter that of oocyte donor age (Figure 3). While Begon et al. (2023) found evidence of a weak effect of sperm provider (paternal) age, we did not. Our finding that the stage of embryo development on transfer was important is supported by previous research which found transfer of a blastocyst was associated with a higher chance of success (Kontopoulos et al., 2019). While previous research on donor cycles has found frozen embryo transfer to be associated with lower live birth rates, and while such a marginal effect is seen in Table 1, this result can be explained in the current cohort by frozen transfers being more likely to occur in those requiring more than one round of embryo transfers, and so occurring in poorer prognosis women, rather than a direct effect of frozen embryos. A potentially unexpected finding was the limited predictive importance of prior autologous ART treatment, with the number of prior autologous transfers only improving discriminatory power by 0.2%.

While previous autologous cycles were of limited importance, the number of previous recipient transfers was one of three key predictive variables, along with oocyte donor age and whether a cleavage stage or blastocyst was transferred (Figure 3 and Supplementary Table A1). Oocyte quality rather than endometrial receptivity has previous been suggested as the reason recipient obesity is not associated with success in oocyte recipient cycles (Jungheim et al., 2013), in contrast to autologous cycles (Sermondade et al., 2019). As use of donor oocytes should results in higher quality oocytes, variation in endometrial receptivity and other patient and treatment factors influencing embryo implantation may explain success or failure. Indeed, previous research has found variation in endometrial thickness to be associated with oocyte recipient outcomes (Noyes et al., 2001).

This research is not without limitations. There were certain variables that may be of importance that were not included in the analysis, such as infertility diagnosis, number of previous miscarriages, the use of pre-implantation genetic testing for aneuploidy (PGT-A) and BMI. Further, our information on the oocytes donor is limited to age and lacks potentially prognostic information such around successful past treatment using their oocytes. However, the source database was updated in 2020 to begin tracking items such as BMI and PGT-A and enable linking of donor and recipient cycles (NPESU, 2019) which will allow future updated versions of the predictive models to be developed and assessed. A key strength of the current analysis is the large sample size. Oocyte donation/recipient cycles make up around 4% of Australian (Newman et al., 2022) and 12% of US ART cycles (Kawwass et al., 2013) which may result in small numbers from individual clinic level analyses, highlighting the importance of national registries as a source of evidence (Hoque et al., 2017). Further, our results provide a baseline for comparison of models utilising expanded features sets or machine learning methods (e.g. random forests) that automatically capture feature interactions not considered in the current analysis. A version of Model 1 can be found online on the YourIVFSuccess website (Figure 4)(YourIVFSuccess, 2023), with Model 3 not appropriate given its use of in-treatment variables (e.g. whether the embryo is cleavage or blastocyst stage).

**Figure 4.**
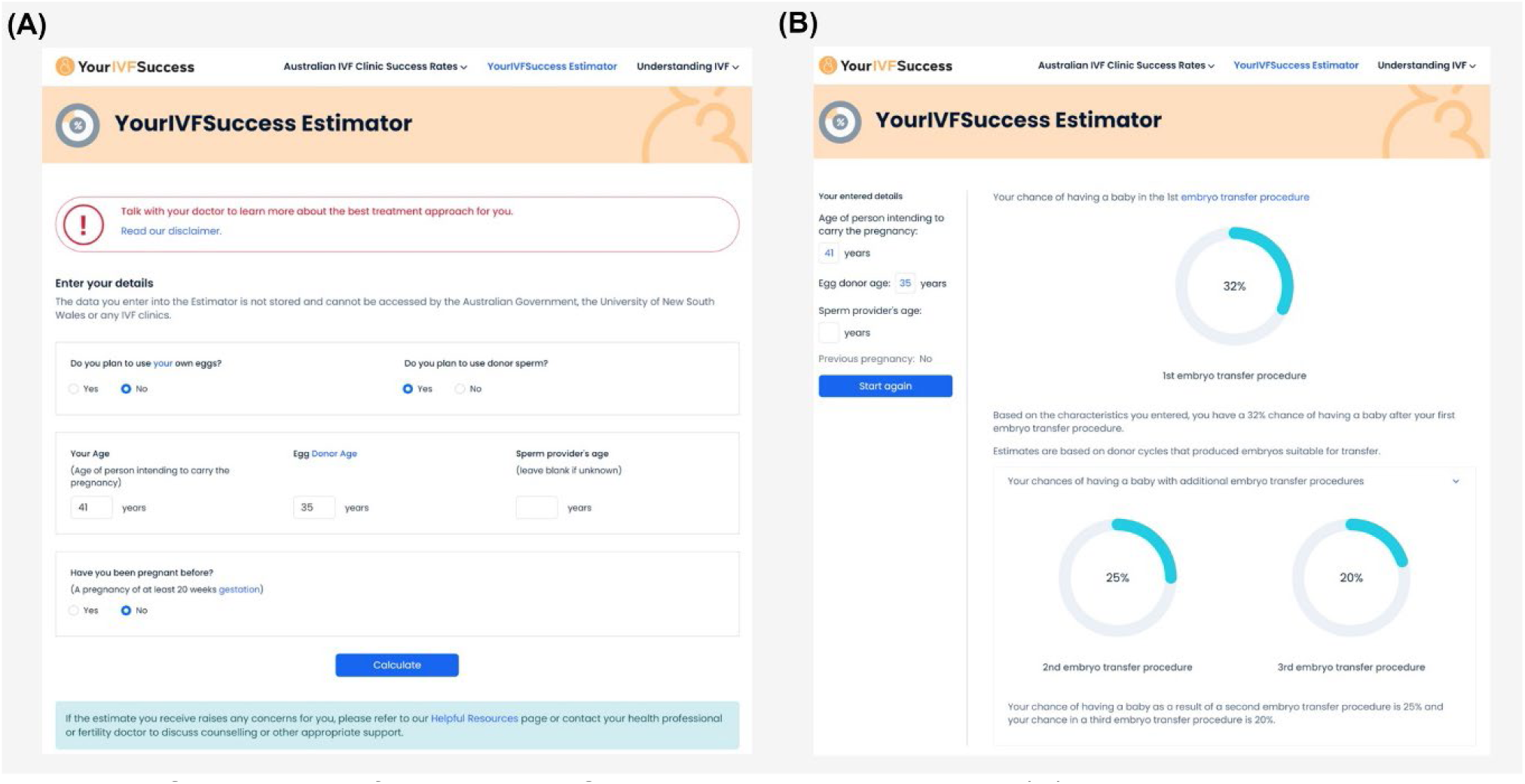
Screenshot of the YourIVFSuccess Estimator showing (A) the webpage where patients enter their data and (B) the predicted live birth rates over the next three embryo transfers based on patient characteristics.

These results confirm the well-established importance of oocyte age as the key patient factor in predicting ART success, while highlighting that in cohorts such as oocyte recipient cycles where oocyte age is rather static it’s value as a predictive factor is reduced. As discussed above the addition of more detailed patient medical histories and physiological measures appears to improve discrimination by around 10%. Given evidence that predictive models help set prospective ART patients expectations improved model quality and importance is of value to patients (Brew et al., 2023), suggesting future research investigate which additional features can be added to maximise this. A related direction of research would be explicitly differentiating between same-sex couple engaging in co-IVF/shared motherhood (Bodri et al., 2018) and the medically necessary use of oocyte donation, which will be feasible using the source database in the future due to a 2020 update (NPESU, 2019).

In many countries patient prognosis is used to determine access to ART funding (Calhaz-Jorge et al., 2020), usually based on crude measures of prognosis such as female age or through restricting the number of funded cycles. While governments rarely use personalised estimates of treatment success, in the commercial sector (particularly the USA) individualised predictions of patient prognosis have increasingly been used to determine access to pooled risk “IVF refund programs” (McLaughlin et al., 2019) (see for example, (Univfy, 2024)).

Concern about funding for unlimited ART treatments in Australia has led to consideration around whether an upper age limit on government funded treatment cycles should be imposed (Keller et al., 2023). However, as shown in the current research, older women maintain a reasonably high chance of a live birth with use of donor oocytes (54.9% of the current cohort achieved a live birth) and this should be considered when maximising resource allocation of healthcare funding while avoiding harm through reducing access to ART of women and couples with reasonably chances of success.

We have shown that clinical prediction models for the likelihood of a live birth after an ART embryo transfer using donated oocytes have reasonable predictive power and were well calibrated to the population of interest. The key variables for prediction of a successful outcome were oocyte donor age, oocyte recipient age, whether the embryo being transferred was cleavage or blastocyst stage and the number of prior recipient embryo transfers. Variable of lessor importance were the oocyte recipient’s parity, the number of prior autologous transfers, the number of embryos being transferred and whether donor or male partner sperm were used. The age of the sperm provider, the use of fresh or frozen-thaw embryos and the use of ICSI or IVF did not appear to impact predictive ability. These prediction models can help an increasingly large proportion of people access ART by aiding in setting the expectations of prospective oocyte recipient ART patients. Future research should investigate which oocyte donor and recipient factors or modelling approaches (e.g. use of machine learning) further improve predictive performance.

## Appendix A Additional Results

**Figure A1.**
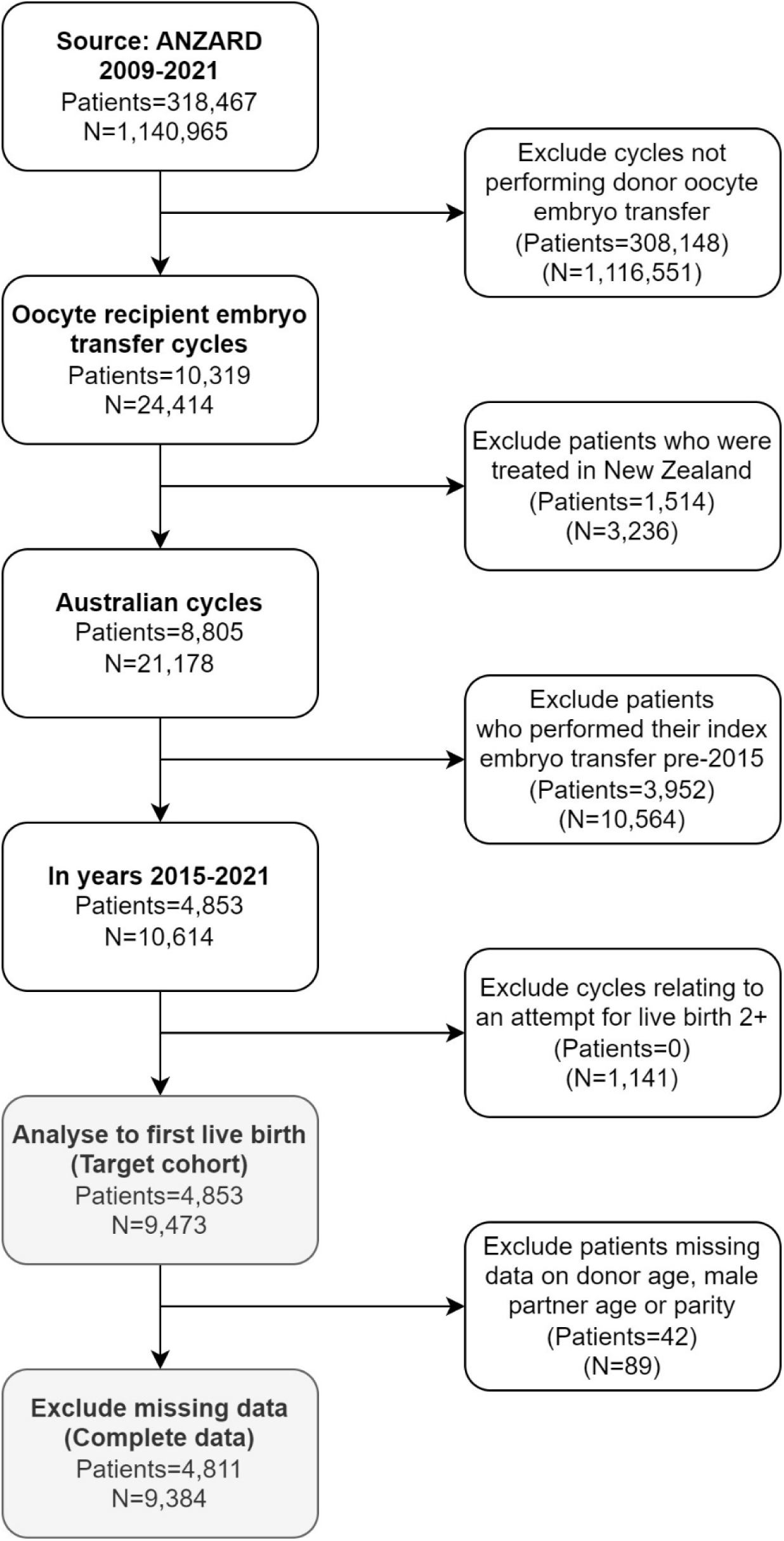
Impact of inclusion and exclusion criteria on the study cohort used to develop an IVF prediction model for donor oocytes. Data sourced from ANZARD (2015-2021). N=number of embryo transfers.

**Figure A2.**
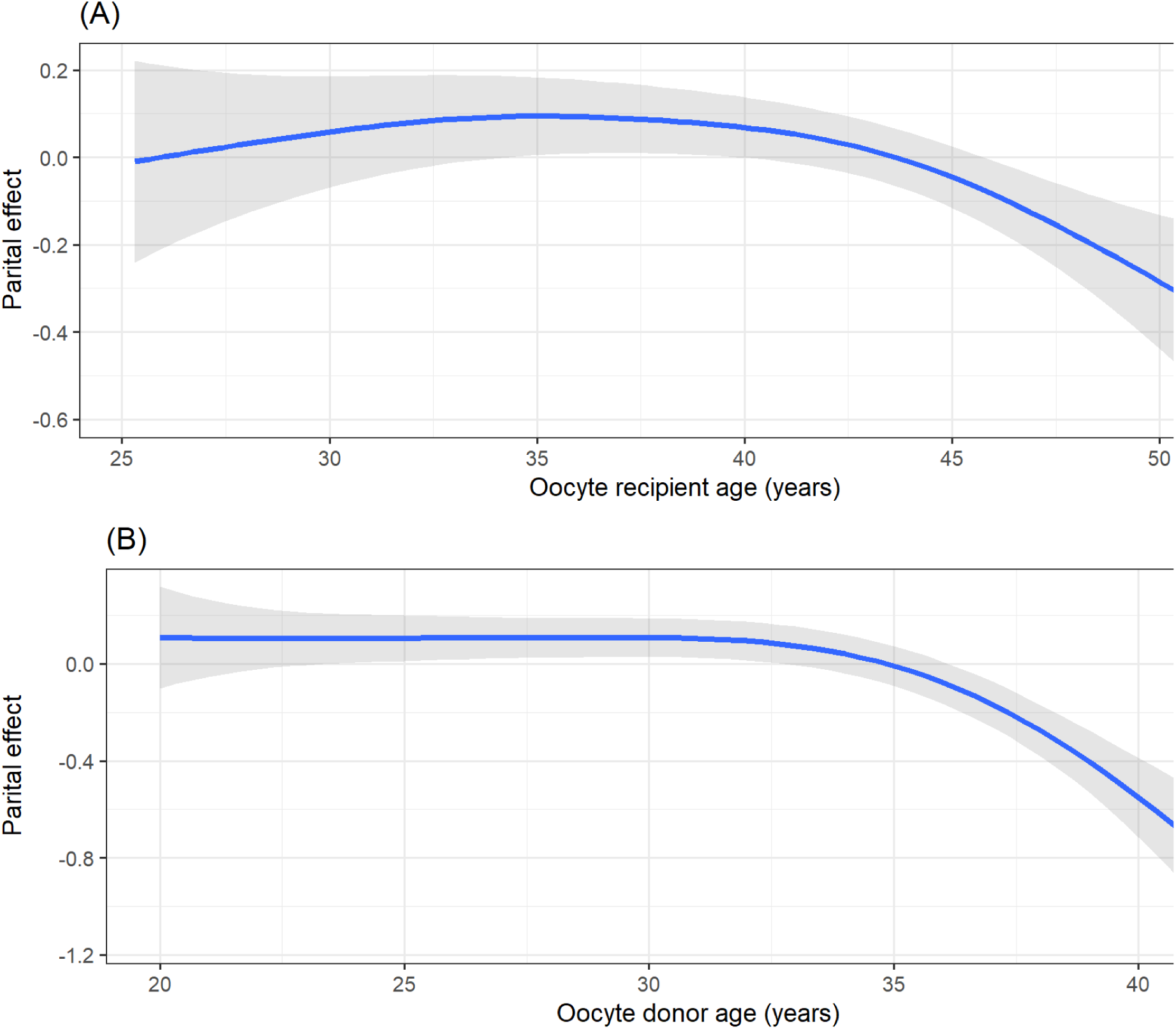
Results from development of an IVF prediction model for donor oocytes using ANZARD (2015-2021). Partial effects after spline transformations from Model 3. (A) Partial effect of oocyte recipient age and (B) Partial effect of oocyte donor age.

**Table A1.** Results from development of an IVF prediction model for donor oocytes using ANZARD (2015-2021). Coefficients of the parametric (non-spline) terms from Model 3.

| Variable | Estimate | Standard error | t-statistic | p value |
| --- | --- | --- | --- | --- |
| Intercept | -0.501 | - | - | - |
| Number of prior oocyte recipient embryo transfers |  |  |  |  |
| Zero | reference |  |  |  |
| One | -0.325 | 0.063 | -5.179 | 0.000 |
| Two or more | -0.528 | 0.065 | -8.148 | 0.000 |
| Parity |  |  |  |  |
| Nulliparous | reference |  |  |  |
| Parous | 0.196 | 0.067 | 2.925 | 0.003 |
| Source of sperm |  |  |  |  |
| Donor | reference |  |  |  |
| Male partner | -0.138 | 0.190 | -0.725 | 0.468 |
| Male partner age | 0.001 | 0.005 | 0.209 | 0.834 |
| Number of prior autologous embryo transfers |  |  |  |  |
| Zero | reference |  |  |  |
| One to three | -0.022 | 0.057 | -0.384 | 0.701 |
| Four or more | -0.110 | 0.061 | -1.815 | 0.070 |
| Number of embryos transferred |  |  |  |  |
| One | reference |  |  |  |
| Two or more | 0.164 | 0.080 | 2.044 | 0.041 |
| Cryopreservation status |  |  |  |  |
| Fresh embryo transfer |  |  |  |  |
| Thaw embryo transfer | -0.056 | 0.055 | -1.009 | 0.313 |
| Embryo development stage |  |  |  |  |
| Cleavage stage embryo transfer | -0.587 | 0.071 | -8.242 | 0.000 |
| Blastocyst | reference |  |  |  |
| Fertilisation method |  |  |  |  |
| IVF | reference |  |  |  |
| Use of ICSI | -0.083 | 0.060 | -1.384 | 0.167 |

## Notes

### Competing Interest Statement

LR declares receipt of consulting fees from Abbott, consulting fees and an educational grant from Merck, role as past president of the Fertility Society of Australia & New Zealand and World Endometriosis Society and a minor shareholding in Monash IVF Group. GC declares receipt of a grant from the Australian Commonwealth Government Medical Research Future Fund Emerging Priorities and Consumer Driven Research Initiative to fund this work.

### Funding Statement

This study was funded by the Australian Commonwealth Government Medical Research Future Fund Emerging Priorities and Consumer Driven Research Initiative

### Author Declarations

Ethics for this project was obtained from the UNSW Sydney Human Research Ethics Committee (reference number: iRECS0859).

